# Elastin degradation markers are elevated in never-smokers with past history of prolonged exposure to secondhand tobacco smoke and are inversely associated with their lung function

**DOI:** 10.1101/2021.12.11.21267498

**Authors:** Jelena Mustra Rakic, Siyang Zeng, Linnea Rohdin-Bibby, Erin L Van Blarigan, Xingjian Liu, Shuren Ma, John P Kane, Rita Redberg, Gerard M. Turino, Eveline Oestreicher Stock, Mehrdad Arjomandi

**Affiliations:** Center for Tobacco Control Research and Education, University of California, San Francisco, California, USA; Cardiovascular Research Institute, University of California, San Francisco, California, USA; Medical Service, San Francisco Veterans Affairs Medical Center; San Francisco; California, USA; Department of Biomedical Informatics and Medical Education, University of Washington, Seattle, Washington, USA; Flight Attendant Medical Research Institute (FAMRI) Bland Lane Center of Excellence on Secondhand Smoke, University of California, San Francisco, California, USA; Department of Epidemiology and Biostatistics, University of California, San Francisco, California, USA; Department of Medicine, Mt Sinai-St Luke’s-Roosevelt Hospital, New York, New York, USA; Division of Cardiology, University of California, San Francisco, California, USA; Division of Pulmonary, Critical Care, Allergy and Immunology, and Sleep Medicine, University of California, San Francisco, California, USA; Division of Occupational and Environmental Medicine; University of California, San Francisco, California, USA

**Keywords:** biomarkers, desmosine/isodesmosine, flight attendants, lung damage, secondhand smoke exposure

## Abstract

**Background:** Prolonged past exposure to secondhand tobacco smoke (SHS) in never-smokers is associated with occult obstructive lung disease and abnormal lung function, in particular reduced diffusing capacity. Previous studies have shown ongoing SHS exposure to be associated with increased elastin degradation markers (EDM) desmosine and isodesmosine.

**Research Question:** Are EDM levels elevated in persons with remote history of SHS exposure, and are those levels associated with reduced lung function?

**Study Design and Methods:** We measured the plasma levels of EDM from 193 never-smoking flight attendants with history of remote but prolonged SHS exposure in aircraft cabin and 103 nonsmoking flight attendants or sea-level control participants without history of cabin SHS exposure, and examined those levels versus their lung function with adjustment for covariates. The cabin SHS exposure was estimated based on airline employment history and dates of smoking ban enactment. EDM plasma levels were quantified by high-performance liquid chromatography and tandem mass spectrometry.

**Results:** The median [interquartile range; IQR] plasma EDM level for all participants was 0.30 [0.24 to 0.36] ng/mL with a total range of 0.16 to 0.65 ng/mL. Plasma EDM levels were elevated in those with history of exposure to cabin SHS compared to those not exposed (0.33±0.08 vs. 0.26±0.06 ng/mL; age- and sex-adjusted P<0.001). In those with history of cabin SHS-exposure, higher EDM levels were associated with lower diffusing capacity (parameter estimate (PE) [95%CI]=4.2 [0.4 to 8.0] %predicted decrease per 0.1 ng/mL increase in EDM; P=0.030). Furthermore, EDM levels were inversely associated with FEV_1_, FEV_1_/FVC, and FEF_25-75_ (PE [95%CI]=5.8 [2.1 to 9.4], 4.0 [2.2 to 5.7], and 12.5 [5.8 to 19.2]% predicted decrease per 0.1 ng/mL increase in EDM, respectively) (P<0.001).

**Interpretation:** Prolonged past exposure to SHS, even when remote, is associated with higher systemic elastin degradation markers that in turn is associated with lower lung function and in particular reduced diffusing capacity.

## BACKGROUND

Secondhand tobacco smoke (SHS), a heterogenous mixture of the sidestream smoke from the burning end of the cigarette and the smoke exhaled by the smoker, is four to 12 times more toxic than the mainstream smoke inhaled by the smoker.^1, 2^ Over the past several decades, a large body of scientific evidence has implicated long-term exposure to SHS as a risk factor for pulmonary diseases including chronic obstructive lung disease (COPD) in both smokers and never-smokers.^3-6^ Remarkably, even remote exposure to secondhand tobacco smoke (SHS) has been shown to be associated with respiratory symptoms and lung function abnormalities consistent with obstructive lung disease.^3, 7-11^

Desmosine and isodesmosine (D/I) are two cross-linked pyridinoline amino acids specific to peptides that are generated from degradation of elastin, a key extracellular protein that provides resilience and elasticity to tissues and is primarily located in lungs, aorta, and skin.^12^ Lung elastin is a recognized target for injury in COPD, and systemic levels of D/I, which are specific for elastin degradation, are elevated in COPD as well as in those without COPD but with acute exposure to tobacco smoke, direct or secondhand.^13-15^ However, it is unclear whether past remote exposure to SHS in those without a diagnosis of COPD is also associated with continued lung injury, elastin degradation, and elevated levels of D/I.

In this study, we aimed to investigate the relationship between remote prolonged exposure to SHS and molecular markers of elastin degradation (plasma D/I) in never-smokers without a diagnosis of COPD, and how those levels are associated with lung function as a way to speculate about the source of elastin degradation product. We hypothesized that there is a positive association between past SHS exposure and plasma D/I, and that the level of plasma D/I is inversely associated with lung function. Presence of such associations would then suggest that past exposure to SHS could result in ongoing lung injury and elastin degradation, contributing to obstructive lung disease.

## STUDY DESIGN AND METHODS

### Study design

To examine our hypothesis above, we took advantage of a “natural experiment” and examined the plasma levels of D/I and lung function from a cohort of healthy flight attendants with no known history of lung diseases including no known COPD who worked for the United States airlines beginning before the smoking ban enactment. These flight attendants were exposed to relatively heavy cabin occupational SHS during their employment for many years and for long periods of time each day.^16^ This set up allowed for a robust objective quantification of cabin SHS exposure using employment history and the dates on which different airlines implemented the smoking bans on domestic and international flights.^16^ Flight attendants who began working after the smoking ban enactment were also recruited as an “unexposed” reference group. Furthermore, blood samples and lung function data from baseline visit of previous participants in MOSES (Multicenter Ozone Study of oldEr Subjects; ClinicalTrials.gov ID# NCT01487005) were also used as additional “unexposed” non-flight attendant reference group.^17^

### Study population

The between June 2014 and October 2019, 241 flight attendants were enrolled as part of an ongoing clinical investigation of the health effects of exposure to cabin SHS in the Flight Attendant Medical Research Institute (FAMRI) Center of Excellence at the University of California San Francisco (UCSF). Thirty-two of those flight attendants had smoked more than 100 cigarettes in their lifetime and were thus excluded from the study. From the 209 remaining participants, 193 had begun their airline employment before the smoking ban enactment and had worked in smoky cabin and were considered “exposed” and 16 were “unexposed”.

All participants gave written informed consent, and the study was approved by the UCSF Institutional Review Board (IRB). Flight attendants were excluded from the study if they smoked more than 100 cigarettes in their lifetime.

The MOSES cohort has been described previously.^17^ Briefly, between 2012 and 2015, 87 healthy nonsmoker adults between the ages of 55 to 70 years old were recruited to participate in a clinical trial investigating the cardiopulmonary health effects of exposure to ambient levels of ozone in a controlled exposure experiments at three centers across the United States. The study consisted of an initial screening visit to determine the eligibility of participants during which blood samples and lung function measurements were collected and incorporated in a biorepository. The data and blood samples from MOSES participants were used in this study to provide a reference “unexposed” group.

All MOSES participants gave written informed consent approved by the respective centers IRB (the University of Rochester Medical Center (URMC), University of North Carolina (UNC), and University of California San Francisco (UCSF)).

### Measurement of cabin SHS exposure

SHS exposure was characterized by a questionnaire developed by UCSF Flight Attendant Medical Research Institute (FAMRI) Center of Excellence,^18^ and modified to acquire information on airline-related occupational history, as described previously.^10, 16^ Briefly, this included employer airlines, duration of employment, and flight routes with quantification of “cabin SHS exposure” as the number of years during which the crewmembers were exposed to SHS in aircraft. Other possible sources of SHS exposure were also explored by questioning participants about their non-cabin exposures in additional settings, as described previously.^19^ Consideration for cabin section was not made.

### Plasma collection and measurements of plasma D/I levels

A non-fasting blood draw at the time of the visit was obtained via forearm venipuncture. The blood samples were collected on ice and centrifuged at 4°C and 1,200 x g for 10 minutes. Plasma was transferred in a new tube and stored at -80°C for further analyses.

Measurement of D/I was done as previously described.^20^ Briefly, plasma samples were acid-hydrolyzed in concentrated hydrochloric acid at 100°C-110°C for 24 hours, and were then applied to a cellulose fiber powders (CF1 or CF11) cartridge to purify. A synthetic desmosine-d4 served as the internal standard for processing plasma samples and measuring D/I. High-performance liquid chromatography and tandem mass spectrometry methods were used for measuring D/I levels.

### Pulmonary function testing

Full pulmonary function testing (PFT) was done for 82 of the eligible participating 209 flight attendants. The remaining 127 participating flight attendants underwent spirometry without plethysmography or diffusing capacity (DCO) measurement.

Full PFT (N=82) were performed in the seated position using a model Vmax 229 CareFusion (CareFusion Corp., Yorba Linda, CA) and nSpire body plethysmograph (nSpire Health Inc., Longmont, CO). This included measurement of the flow-volume curve and spirometry;^21^ lung volume by single breath dilution;^22, 23^ and plethysmography;^24^ airway resistance during panting at functional residual capacity (FRC);^25, 26^ and single breath carbon monoxide diffusing capacity.^27^ Spirometry without plethysmography or diffusing capacity measurement for the 127 flight attendants was done using a portable spirometer (EasyOne, NDD Medical Technologies) in the seated position. MOSES lung function testing procedures have been described previously.^17^ Briefly, spirometry was performed in seated position using a dry rolling seal spirometer calibrated weekly: URMC used a KoKo PFT Spirometer (Nspirehealth, Longmont, CO); UNC used VIASYS 10.2-L model 1022 (SensorMedics; Palm Springs, CA); and UCSF employed an S&M Instrument, PDS Instrumentation (Louisville, CA).

All pulmonary function studies were conducted according to the American Thoracic Society (ATS) and European Respiratory Society (ERS) guidelines.^28-33^ Participants did not undergo bronchodilator administration. The Global Lung Initiative (GLI) predicted formulas were used to compute the percent predicted values as well as lower and upper limit of normal values for spirometry measures (FEV_1_/FVC, FEV_1_, FVC, FEF_25-75_).^34^ Crapo predicted formulas were used to compute the percent predicted values as well as lower and upper limits of normal values for diffusing capacity.^35, 36^ Spiromteric COPD was defined using Global Initiative for Obstructive Lung Disease (GOLD) criteria unless otherwise specified.^37^

### Statistical analysis

Participants’ characteristics including demographics, years of SHS exposure, and lung function measures were examined and summarized within all participants and with respect to subgroups with or without cabin SHS exposure. The adjusted plasma D/I levels were computed by calculating the residual values of the raw plasma D/I levels and their predicted values from a linear regression model of the plasma D/I levels over age, sex, height, and weight. A comparison of the distributions was performed using an unpaired t-test for each continuous variable or a Chi-squared test for each binary or categorical variable. The P values and the descriptive statistics including the mean ± standard deviation (SD), median [1^st^ quartile, 3^rd^ quantile] for continuous variables or N (%) for binary and categorical variables were presented.

The associations between plasma D/I levels and lung function measures were examined, in the whole group of participants and the subgroup of those who had cabin SHS exposure, using linear regression modelling with adjustment for covariates including age, sex, height, and weight. The associations between having history of cabin SHS exposure as well as years of cabin SHS exposure and lung function measures were examined using linear regression modelling with adjustment for the same covariates. For each individual model using one of the lung volume measures as the dependent variable, the total number of participants involved in the model and the parameter estimate with a 95% confidence interval and a P value for plasma D/I levels or years of SHS exposure were reported accordingly.

The associations between plasma D/I levels and years of SHS exposure were examined using linear regression modelling with adjustments for the same covariates in the subgroup of those who had cabin SHS exposure. The difference in plasm D/I levels between the subgroups with and without cabin SHS exposure was assessed using linear regression modeling with adjustments for the same covariates in the whole group of participants. For these models using plasma D/I levels as the dependent variable, the total number of participants involved in the model and the parameter estimate with a 95% confidence interval and a P-value for years of SHS exposure or the binary indicator of having past SHS exposure were reported accordingly.

Statistical analyses were conducted using the R (version 3.6) statistical software. A significance level of α<0.05 was used to determined statistical significance.

## RESULTS

### Participants’ characteristics

From the total of 241 flight attendants who were initially recruited into the study, 32 were excluded because of they were not never-smokers. Among the remaining 209, 193 (92.3%) had been exposed to cabin SHS and 16 had not been exposed to cabin SHS. Additionally, 87 non-flight attendants healthy nonsmoking participants (from MOSES) were included in the SHS unexposed group. Overall, 296 participants were included in the analyses (**Figure 1**) consisting of 193 (65.2%) exposed to cabin SHS and 103 (34.8%) unexposed.

**Figure 1.**
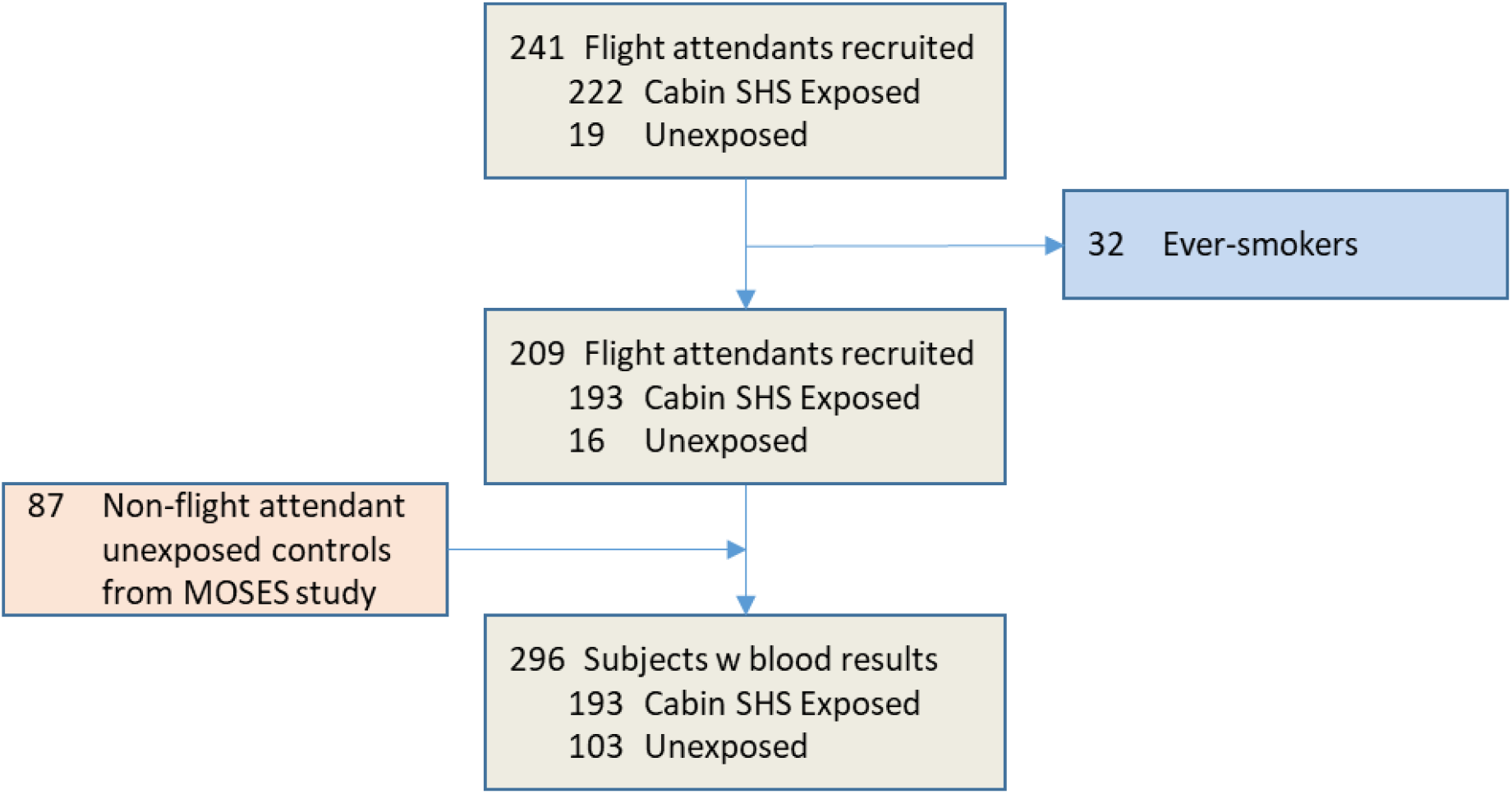
Participants’ flowchart. The flow of participants through the study screening and procedures.

Participants’ characteristics are shown in **Table 1**. The average age of participants (N=296) was 64.0±7.8 years. The SHS unexposed group were younger (59.3±6.0 years) than the SHS exposed group (66.5±7.4 years) with majority being women in both groups (63% and 81%, respectively). The cohort was primarily composed of people of White racial background (85.5%), but also included people who identified their racial background as Asian, African American, Indian and Alaskan Native, Native Hawaiian or other Pacific Islander. There was no significant race/ethnicity difference between SHS exposed and unexposed groups. The average body mass index (BMI) was 24.4±3.5 Kg/m^2^ (21 participants were obese defined by BMI >30 Kg/m^2^), with no significant difference in BMI between SHS exposed and unexposed groups. Amongst all participants, the total years of airline employment and cabin SHS exposure were 31.1±11.4 and 17.8±9.4 years, respectively.

**Table 1.**
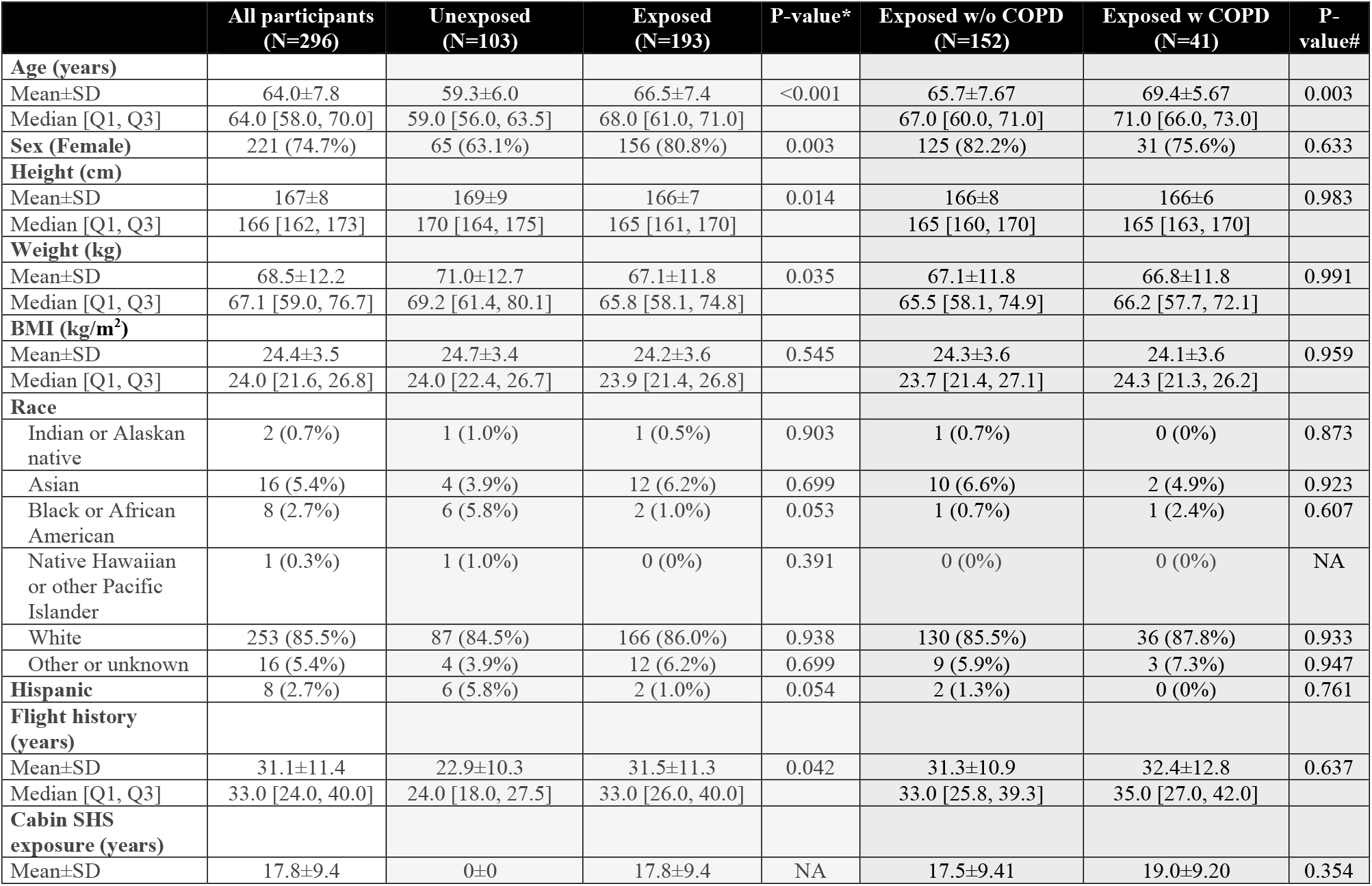
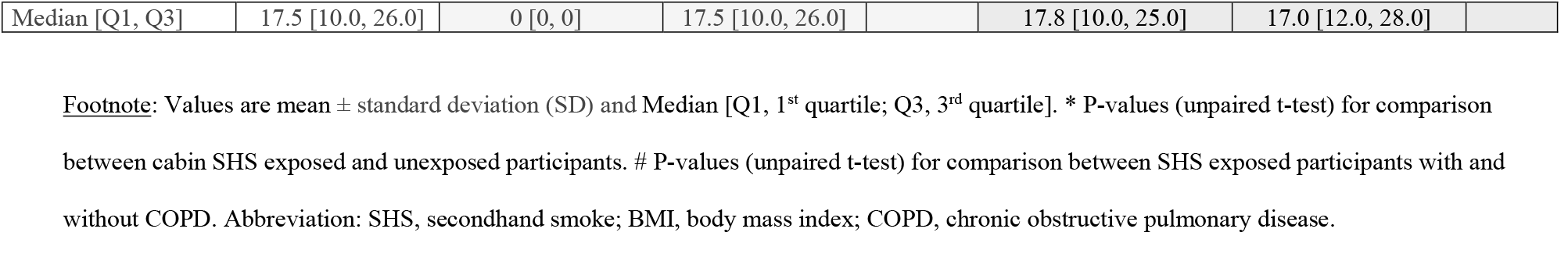
Participant characteristics.

|  | All participants<br>(N=296) | Unexposed<br>(N=103) | Exposed<br>(N=193) | P-value* | Exposed w/o COPD<br>(N=152) | Exposed w COPD<br>(N=41) | P-value# |
| --- | --- | --- | --- | --- | --- | --- | --- |
| **Age (years)** |  |  |  |  |  |  |  |
| Mean±SD | 64.0±7.8 | 59.3±6.0 | 66.5±7.4 | <0.001 | 65.7±7.67 | 69.4±5.67 | 0.003 |
| Median [Q1, Q3] | 64.0 [58.0, 70.0] | 59.0 [56.0, 63.5] | 68.0 [61.0, 71.0] |  | 67.0 [60.0, 71.0] | 71.0 [66.0, 73.0] |  |
| **Sex (Female)** | 221 (74.7%) | 65 (63.1%) | 156 (80.8%) | 0.003 | 125 (82.2%) | 31 (75.6%) | 0.633 |
| **Height (cm)** |  |  |  |  |  |  |  |
| Mean±SD | 167±8 | 169±9 | 166±7 | 0.014 | 166±8 | 166±6 | 0.983 |
| Median [Q1, Q3] | 166 [162, 173] | 170 [164, 175] | 165 [161, 170] |  | 165 [160, 170] | 165 [163, 170] |  |
| **Weight (kg)** |  |  |  |  |  |  |  |
| Mean±SD | 68.5±12.2 | 71.0±12.7 | 67.1±11.8 | 0.035 | 67.1±11.8 | 66.8±11.8 | 0.991 |
| Median [Q1, Q3] | 67.1 [59.0, 76.7] | 69.2 [61.4, 80.1] | 65.8 [58.1, 74.8] |  | 65.5 [58.1, 74.9] | 66.2 [57.7, 72.1] |  |
| **BMI (kg/m<sup>2</sup>)** |  |  |  |  |  |  |  |
| Mean±SD | 24.4±3.5 | 24.7±3.4 | 24.2±3.6 | 0.545 | 24.3±3.6 | 24.1±3.6 | 0.959 |
| Median [Q1, Q3] | 24.0 [21.6, 26.8] | 24.0 [22.4, 26.7] | 23.9 [21.4, 26.8] |  | 23.7 [21.4, 27.1] | 24.3 [21.3, 26.2] |  |
| **Race** |  |  |  |  |  |  |  |
| Indian or Alaskan native | 2 (0.7%) | 1 (1.0%) | 1 (0.5%) | 0.903 | 1 (0.7%) | 0 (0%) | 0.873 |
| Asian | 16 (5.4%) | 4 (3.9%) | 12 (6.2%) | 0.699 | 10 (6.6%) | 2 (4.9%) | 0.923 |
| Black or African American | 8 (2.7%) | 6 (5.8%) | 2 (1.0%) | 0.053 | 1 (0.7%) | 1 (2.4%) | 0.607 |
| Native Hawaiian or other Pacific Islander | 1 (0.3%) | 1 (1.0%) | 0 (0%) | 0.391 | 0 (0%) | 0 (0%) | NA |
| White | 253 (85.5%) | 87 (84.5%) | 166 (86.0%) | 0.938 | 130 (85.5%) | 36 (87.8%) | 0.933 |
| Other or unknown | 16 (5.4%) | 4 (3.9%) | 12 (6.2%) | 0.699 | 9 (5.9%) | 3 (7.3%) | 0.947 |
| **Hispanic** | 8 (2.7%) | 6 (5.8%) | 2 (1.0%) | 0.054 | 2 (1.3%) | 0 (0%) | 0.761 |
| **Flight history (years)** |  |  |  |  |  |  |  |
| Mean±SD | 31.1±11.4 | 22.9±10.3 | 31.5±11.3 | 0.042 | 31.3±10.9 | 32.4±12.8 | 0.637 |
| Median [Q1, Q3] | 33.0 [24.0, 40.0] | 24.0 [18.0, 27.5] | 33.0 [26.0, 40.0] |  | 33.0 [25.8, 39.3] | 35.0 [27.0, 42.0] |  |
| **Cabin SHS exposure (years)** |  |  |  |  |  |  |  |
| Mean±SD | 17.8±9.4 | 0±0 | 17.8±9.4 | NA | 17.5±9.41 | 19.0±9.20 | 0.354 |
| Median [Q1, Q3] | 17.5 [10.0, 26.0] | 0 [0, 0] | 17.5 [10.0, 26.0] |  | 17.8 [10.0, 25.0] | 17.0 [12.0, 28.0] |  |

Footnote: Values are mean ± standard deviation (SD) and Median [Q1, 1<sup>st</sup> quartile; Q3, 3<sup>rd</sup> quartile]. \* P-values (unpaired t-test) for comparison between cabin SHS exposed and unexposed participants. # P-values (unpaired t-test) for comparison between SHS exposed participants with and without COPD. Abbreviation: SHS, secondhand smoke; BMI, body mass index; COPD, chronic obstructive pulmonary disease.

### Lung function measurements

Pulmonary function test measurements are shown in **Table 2**. Although none of the participants had clinical diagnosis of COPD, 53 (17.9%) and 22 (7.4%) had abnormal FEV_1_ to FVC ratio consistent with spirometric COPD by Global Initiative for Obstructive Lung Disease (GOLD) (FEV1/FVC <0.70) and the lower limit of normal (LLN) criteria, respectively (FEV_1_/FVC=0.74±0.07 at 94±9% predicted). Among the SHS exposed group, 41 (21.2%) and 14 (7.3%) participants had abnormal FEV_1_/FVC consistent with spirometric COPD by GOLD and LLN criteria, respectively. Among the SHS unexposed group, 12 (11.7%) participants had FEV_1_/FVC <0.70, but only 8 (7.8%) had spirometric COPD by LLN criteria. The FEV_1_ was within the normal range for both unexposed and exposed participants as a whole, but was reduced at 1.91±0.55 L (79±20 %predicted) in the SHS exposed participants with spirometric COPD.

**Table 2.** Pulmonary function test and elastin degradation product measurements.

|  | All participants<br>(N=296) | Unexposed<br>(N=103) | Exposed<br>(N=193) | P-value* | Exposed w/o COPD<br>(N=152) | Exposed w COPD<br>(N=41) | P-value# |
| --- | --- | --- | --- | --- | --- | --- | --- |
| **FEV<sub>1</sub> (L)** |  |  |  |  |  |  |  |
| Mean±SD | 2.55±0.68 | 2.93±0.62 | 2.36±0.62 | <0.001 | 2.48±0.589 | 1.91±0.548 | <0.001 |
| Median [Q1, Q3] | 2.43 [2.10, 2.93] | 2.79 [2.50, 3.35] | 2.24 [1.97, 2.60] |  | 2.36 [2.11, 2.77] | 1.92 [1.72, 2.16] |  |
| **FEV<sub>1</sub> (% predicted)** |  |  |  |  |  |  |  |
| Mean±SD | 97±18 | 101±14.0 | 95±20 | 0.004 | 99.1±17.3 | 78.9±19.5 | <0.001 |
| Median [Q1, Q3] | 98 [86, 107] | 101 [91, 110] | 95 [84, 106] |  | 97.8 [87.4, 107] | 77.7 [69.9, 90.6] |  |
| **FVC (L)** |  |  |  |  |  |  |  |
| Mean±SD | 3.44±0.85 | 3.89±0.84 | 3.20±0.76 | <0.001 | 3.26±0.770 | 3.00±0.677 | 0.116 |
| Median [Q1, Q3] | 3.25 [2.84, 3.90] | 3.77 [3.17, 4.52] | 3.05 [2.72, 3.49] |  | 3.08 [2.75, 3.56] | 2.98 [2.54, 3.27] |  |
| **FVC (% predicted)** |  |  |  |  |  |  |  |
| Mean±SD | 102±16 | 105±14 | 100±17 | 0.023 | 102±17.0 | 95.8±16.9 | 0.157 |
| Median [Q1, Q3] | 101 [91.3, 112] | 104 [96.3, 115] | 99.0 [89.8, 111] |  | 100 [90.0, 111] | 95.2 [81.6, 110] |  |
| **FEV<sub>1</sub>/FVC** |  |  |  |  |  |  |  |
| Mean±SD | 0.74±0.07 | 0.76±0.05 | 0.73±0.08 | 0.012 | 0.76±0.03 | 0.63±0.09 | <0.001 |
| Median [Q1, Q3] | 0.75 [0.71, 0.78] | 0.75 [0.72, 0.79] | 0.74 [0.70, 0.78] |  | 0.76 [0.73, 0.79] | 0.66 [0.61, 0.68] |  |
| **FEV<sub>1</sub>/FVC < 0.70** | 53 (17.9%) | 12 (11.7%) | 41 (21.2%) | 0.140 | 0 (0%) | 41 (100%) | NA |
| **FEV<sub>1</sub>/FVC (% predicted)** |  |  |  |  |  |  |  |
| Mean±SD | 94±9 | 96±6 | 94±10 | 0.064 | 97.0±4.96 | 80.9±11.6 | <0.001 |
| Median [Q1, Q3] | 95.0 [90.3, 99.4] | 95.4 [92.5, 99.9] | 94.7 [89.6, 99.3] |  | 97.1 [93.2, 100] | 84.9 [78.8, 87.5] |  |
| **Peak expiratory flow (L/s)** |  |  |  |  |  |  |  |
| Mean±SD | 368±118 | 395±85.0 | 366±120 | 0.668 | 386±122 | 310±94.6 | 0.001 |
| Median [Q1, Q3] | 343 [300, 409] | 390 [343, 450] | 342 [297, 406] |  | 355 [311, 419] | 309 [259, 333] |  |
| **FEF<sub>25-75</sub> (L/s)** |  |  |  |  |  |  |  |
| Mean±SD | 2.05±0.84 | 2.43±0.77 | 1.85±0.81 | <0.001 | 2.10±0.713 | 0.958±0.390 | <0.001 |
| Median [Q1, Q3] | 1.97 [1.45, 2.58] | 2.31 [1.84, 2.97] | 1.78 [1.30, 2.32] |  | 1.98 [1.58, 2.56] | 0.960 [0.670, 1.26] |  |
| **FEF<sub>25-75</sub> (% predicted)** |  |  |  |  |  |  |  |
| Mean±SD | 890±32 | 95±27 | 87±35 | 0.061 | 97.1±30.0 | 47.7±19.2 | <0.001 |
| Median [Q1, Q3] | 88.3 [68.7, 110] | 92.3 [75.7, 110] | 83.8 [64.6, 105] |  | 93.8 [73.0, 113] | 48.4 [34.1, 63.7] |  |
| **DCO (mL/min/mmHg)** |  |  |  |  |  |  |  |
| Mean±SD | 20.5±4.2 | 23.1±3.8 | 20.2±4.1 | 0.14 | 20.5±4.23 | 18.6±2.88 | 0.172 |
| Median [Q1, Q3] | 20.2 [17.4, 22.9] | 23.2 [20.8, 25.6] | 19.9 [17.3, 22.2] |  | 20.2 [17.4, 22.5] | 18.6 [17.3, 19.7] |  |
| **DCO (% predicted)** |  |  |  |  |  |  |  |
| Mean±SD | 80±12 | 84±12 | 80±12 | 0.576 | 80.5±11.8 | 74.9±9.70 | 0.254 |
| Median [Q1, Q3] | 80 [71, 89] | 84 [75, 92] | 79 [71, 88] |  | 81.1 [71.9, 89.8] | 75.0 [68.1, 80.2] |  |
| **D/I (ng/mL)** |  |  |  |  |  |  |  |
| Mean±SD | 0.31±0.08 | 0.26±0.06 | 0.33±0.08 | <0.001 | 0.319±0.0775 | 0.378±0.0832 | <0.001 |
| Median [Q1, Q3] | 0.30 [0.24, 0.36] | 0.24 [0.22, 0.29] | 0.33 [0.27, 0.37] |  | 0.310 [0.264, 0.358] | 0.367 [0.340, 0.414] |  |
| **Adjusted D/I (normalized score)** |  |  |  |  |  |  |  |
| Mean±SD | 0±0.07 | -0.02±0.06 | 0.01±0.07 | 0.002 | 0.002±0.07 | 0.04±0.07 | 0.020 |
| Median [Q1, Q3] | -0.01 [-0.05, 0.04] | -0.02 [-0.06, 0.01] | 0.001 [-0.04, 0.05] |  | -0.003 [-0.04, 0.04] | 0.02 [-0.01, 0.09] |  |

Footnote: Values are mean ± standard deviation (SD) and Median [Q1, 1<sup>st</sup> quartile; Q3, 3<sup>rd</sup> quartile]. \* P-values (unpaired t-test) for comparison between cabin SHS exposed and unexposed participants. # P-values (unpaired t-test) for comparison between SHS exposed participants with and without COPD. Abbreviation: SHS, secondhand smoke; FEV<sub>1</sub>, forced expiratory volume in 1 second; FVC, forced vital capacity; FEF, forced expiratory flow; FET, forced expiratory time; DCO, diffusing capacity; D/I, desmosine/isodesmosine.

The diffusion capacity, which was measured only in 82 participants (all flight attendants), was (20.5±4.2 mL/min/mmHg at 80±12 %predicted), and was below the LLN in 32 (39%) of participants. Although not statistically significant, the diffusion capacity of the SHS exposed group was lower than that of the SHS unexposed group (20.2±4.1 versus 23.1±3.8 mL/min/mmHg [80±12 versus 84±12%predicted]). Similarly, the diffusing capacity of SHS exposed group with spirometric COPD was non-significantly lower than that of the SHS exposed group without spirometric COPD (18.6±2.9 versus 20.5±4.2 mL/min/mmHg [75±10 versus 81±12 %predicted]).

### Associations of plasma levels of elastin degradation products with lung function measures

In linear regression models adjusted for age, sex, height and weight, plasma levels of elastin degradation products (desmosine and isodesmosine [D/I]) were inversely associated with FEV_1_, FVC, FEV_1_/FVC, FEF_25-75_, and DCO among all participants (FEV_1_: ß=-1.76L, P<0.001; FVC: ß=-1.43 L, P=0.002; FEV_1_/FVC: ß=-26%, P<0.001; FEF_25-_75: ß=-2.74L, P<0.001; and DCO: ß=-9.98 mL/min/mmHg, P=0.037) (**Table 3**). Similar associations were found with the percent predicted values (**Figure 2**).

**Table 3.**
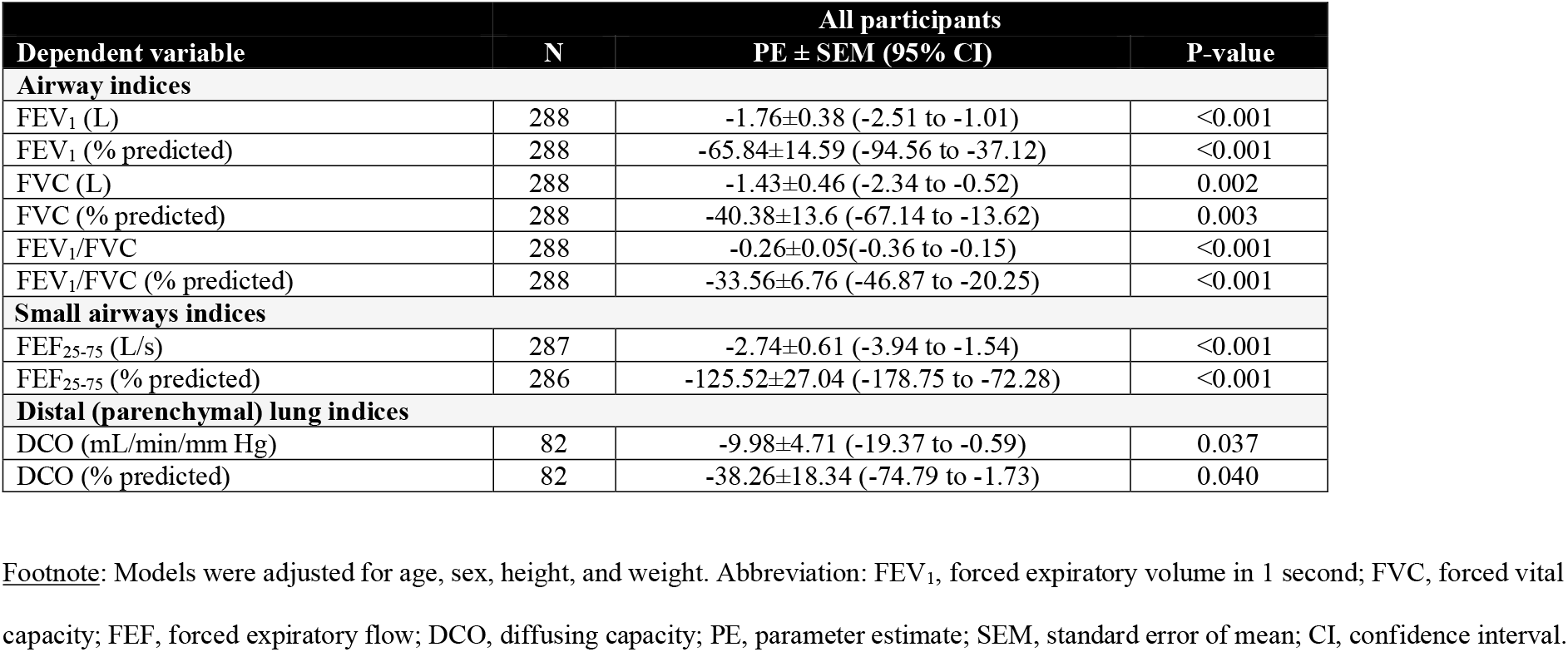
Associations between plasma levels of elastin degradation products (desmosine and isodesmosine, D/I) and lung function measures.

| **Dependent variable** | **N** | **All participants**<br>**PE ± SEM (95% CI)** | **P-value** |
| --- | --- | --- | --- |
| **Airway indices** |  |  |  |
| FEV<sub>1</sub> (L) | 288 | -1.76±0.38 (-2.51 to -1.01) | <0.001 |
| FEV<sub>1</sub> (% predicted) | 288 | -65.84±14.59 (-94.56 to -37.12) | <0.001 |
| FVC (L) | 288 | -1.43±0.46 (-2.34 to -0.52) | 0.002 |
| FVC (% predicted) | 288 | -40.38±13.6 (-67.14 to -13.62) | 0.003 |
| FEV<sub>1</sub>/FVC | 288 | -0.26±0.05(-0.36 to -0.15) | <0.001 |
| FEV<sub>1</sub>/FVC (% predicted) | 288 | -33.56±6.76 (-46.87 to -20.25) | <0.001 |
| **Small airways indices** |  |  |  |
| FEF<sub>25-75</sub> (L/s) | 287 | -2.74±0.61 (-3.94 to -1.54) | <0.001 |
| FEF<sub>25-75</sub> (% predicted) | 286 | -125.52±27.04 (-178.75 to -72.28) | <0.001 |
| **Distal (parenchymal) lung indices** |  |  |  |
| DCO (mL/min/mm Hg) | 82 | -9.98±4.71 (-19.37 to -0.59) | 0.037 |
| DCO (% predicted) | 82 | -38.26±18.34 (-74.79 to -1.73) | 0.040 |

Footnote: Models were adjusted for age, sex, height, and weight. Abbreviation: FEV<sub>1</sub>, forced expiratory volume in 1 second; FVC, forced vital capacity; FEF, forced expiratory flow; DCO, diffusing capacity; PE, parameter estimate; SEM, standard error of mean; CI, confidence interval.

**Figure 2.**
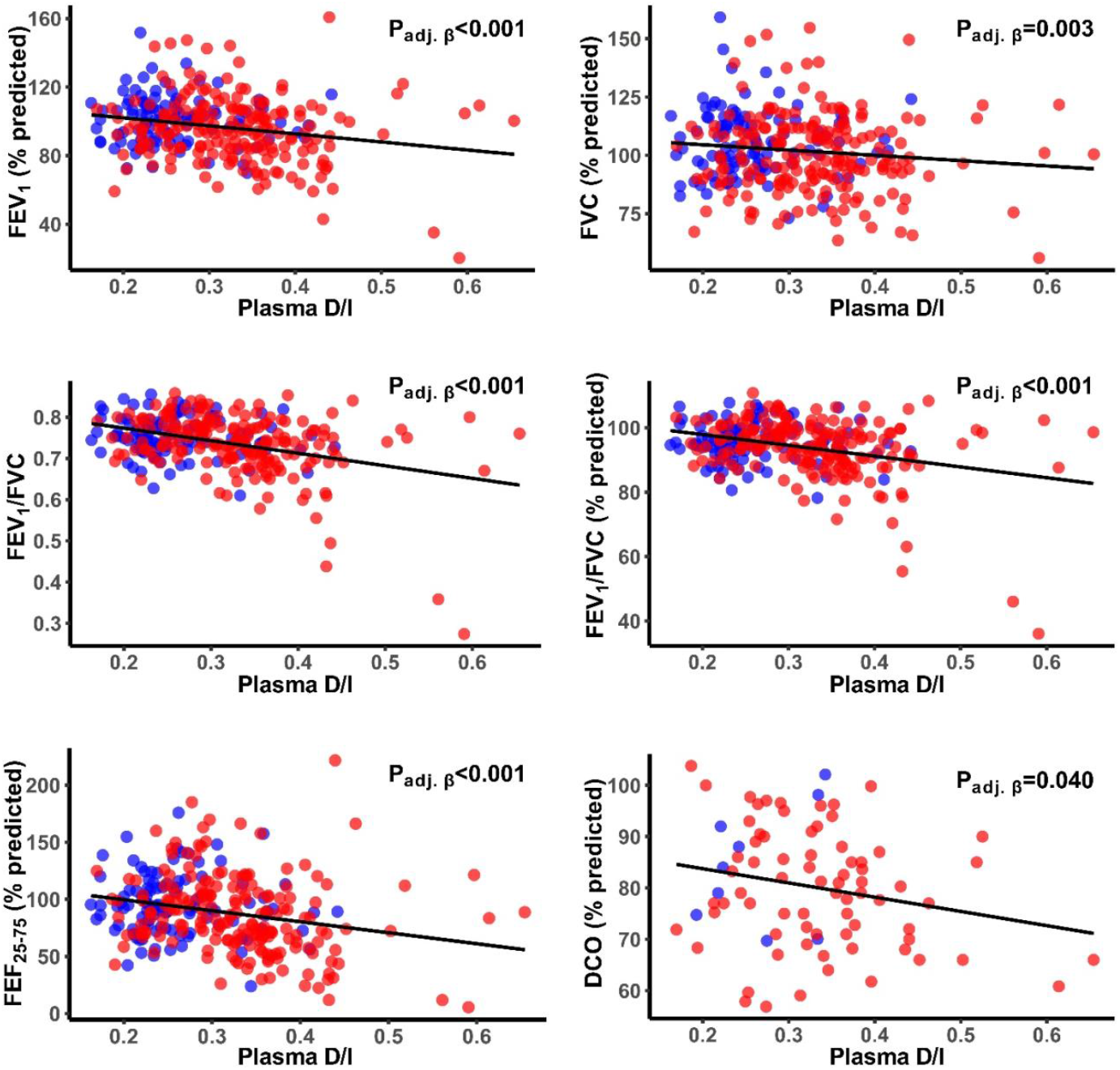
Association of Systemic levels of elastin degradation products (plasma desmosine and isodesmosine [D/I]) with lung function. Scatter plots representing the association of adjusted plasma levels of elastin degradation products (D/I) with lung function among cabin SHS-exposed participants. P-values (P_adj. β_) were obtained from multivariate regression models with adjustment for age, sex, height, and weight. Blue and red dots represent SHS-unexposed and SHS-exposed participants, respectively.

Among the subgroup with cabin SHS-exposure, we found similar inverse association between plasma levels of D/I and FEV_1_, FEV_1_/FVC, FEF_25-75_, and DCO in adjusted models (FEV_1_: ß=-1.47L, P=0.001; FEV_1_/FVC: ß=-31%, P<0.001; FEF_25-75_: ß=-2.55L, P<0.001; and DCO: ß=-10.92 mL/min/mm Hg, P=0.025; respectively) (**Table S1**). Similar associations were found with the percent predicted values (**Figure 2**). When SHS-exposed group was divided into the subgroups with and without COPD, similar directions of the associations were observed, although they did not reach statistical significance **(Table S1**).

### Associations of SHS exposure with plasma levels of elastin degradation products

Among all participants, the unadjusted and adjusted (age, sex, height, and weight-adjusted) plasma levels of D/I were significantly elevated in the SHS-exposed group compared to the SHS-unexposed group (unadjusted levels: 0.33±0.08 vs. 0.26±0.06 ng/mL, P<0.001 (**Table 2**); adjusted levels in **Figure 3**). Among SHS-exposed group, those with spirometric COPD had significantly elevated levels of plasma D/I compared to those without spirometric COPD (unadjusted levels: 0.38±0.08 vs. 0.32±0.08 ng/mL, P<0.001 (**Table 2**); adjusted levels in **(Figure 3)**.

**Figure 3.**
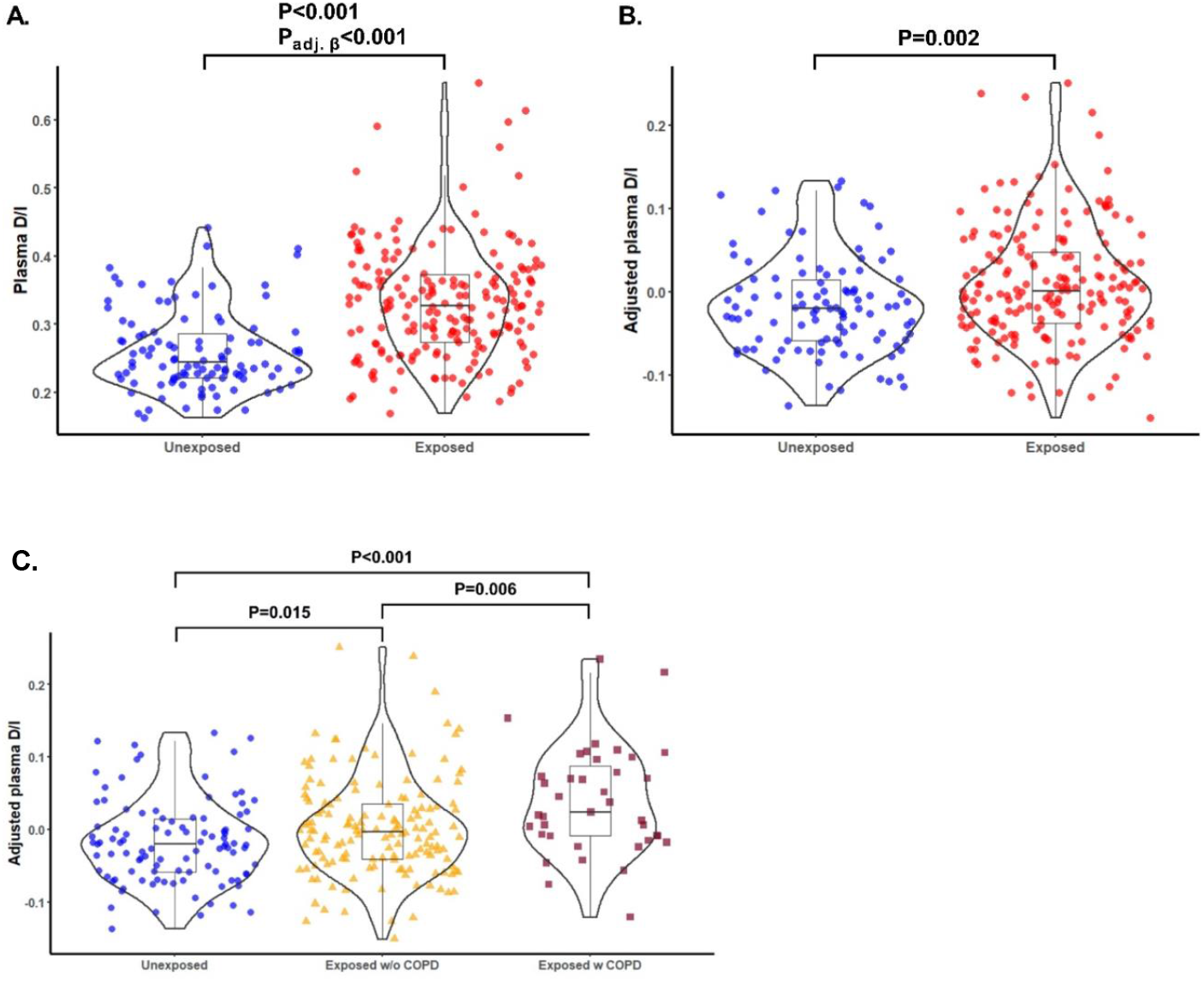
Systemic levels of elastin degradation products (plasma desmosine and isodesmosine) in SHS-exposed and unexposed participants (with and without spirometric COPD). Plasma levels of elastin degradation products (D/I) from exposed and unexposed participants without and with adjustments are plotted using violin plots (**A** and **B**, respectively). Plasma levels of D/I from exposed and unexposed participants without and with COPD are plotted using violin plots (**C**). Adjusted plasma levels of D/I were obtained by computing the residuals from regression modeling with adjustment for age, sex, height, and weight. P-values were obtained from both pairwise t-test comparisons (P) and multivariate regression models with adjustment for age, sex, height, and weight (P_adj. β_).

In the univariate model, plasma levels of D/I were directly associated with years of SHS exposure, and with age, but not with length of employment as flight attendants (P<0.001, P<0.001, and P=0.43, respectively) (**Table S2**). However, in the multivariate model, after adjusting for age, sex, height, and weight; years of cabin SHS exposure and total years of flight employment were not associated with plasma D/I levels (**Table S2**).

### Associations of SHS exposure with lung function measures

Among all participants, history of exposure to cabin SHS was inversely associated with FEV_1_, FVC, and FEF_25-75_ in models adjusted for age, sex, height, and weight (FEV_1_: ß=-0.25L, P<0.001; FVC: ß=-0.30L, P<0.001; and FEF_25-75_: ß=-0.23L, P=0.020) (**Table 4**). As an example, those exposed to cabin SHS smoke had an FEV_1_ that was 247 mL lower compared to those who were not exposed to cabin SHS. The associations between exposure to SHS and FEV_1_/FVC or DCO were in the hypothesized directions but did not reach statistical significance.

**Table 4.** Associations between cabin SHS exposure and lung function measures.

| Dependent variables | All participants<br>Independent variable: Exposed (Y/N) |  |  | All participants<br>Independent variable: Years of exposure |  |  |
| --- | --- | --- | --- | --- | --- | --- |
|  | N | PE±SEM<br>(95% CI) | P-value | N | PE±SEM<br>(95% CI) | P-value |
| **Airway indices** |  |  |  |  |  |  |
| FEV<sub>1</sub> (L) | 288 | -0.247±0.061<br>(-0.368 to -0.126) | <0.001 | 288 | -0.004±0.003<br>(-0.010 to 0.0014) | 0.140 |
| FEV<sub>1</sub> (% predicted) | 288 | -8.923±2.349<br>(-13.547 to -4.298) | <0.001 | 288 | -0.14±0.11<br>(-0.37 to 0.08) | 0.198 |
| FVC (L) | 288 | -0.303±0.073<br>(-0.446 to -0.160) | <0.001 | 288 | -0.006±0.004<br>(-0.013 to 0.001) | 0.108 |
| FVC (% predicted) | 288 | -8.200±2.146<br>(-12.424 to -3.976) | <0.001 | 288 | -0.13±0.10<br>(-0.33 to 0.07) | 0.198 |
| FEV<sub>1</sub>/FVC (%) | 288 | -0.01±0.01<br>(-0.03 to 0.01) | 0.311 | 288 | 0.0001±0.0004<br>(-0.0009 to 0.0007) | 0.798 |
| FEV<sub>1</sub>/FVC (% predicted) | 288 | -1.30±1.12<br>(-3.51 to 0.90) | 0.246 | 288 | -0.02±0.05<br>(-0.12 to 0.09) | 0.766 |
| **Small airways indices** |  |  |  |  |  |  |
| FEF<sub>25-75</sub> (L/s) | 287 | -0.230±0.099<br>(-0.424 to -0.036) | 0.020 | 287 | -0.005±0.005<br>(-0.015 to 0.004) | 0.268 |
| FEF<sub>25-75</sub> (% predicted) | 286 | -10.052±4.388<br>(-18.690 to -1.414) | 0.022 | 286 | -0.27±0.21<br>(-0.68 to 0.15) | 0.205 |
| **Distal (parenchymal) lung indices** |  |  |  |  |  |  |
| DCO (mL/min/mm Hg) | 82 | -1.206±1.185<br>(-3.566 to 1.153) | 0.311 | 82 | -0.004±0.037<br>(-0.079 to 0.070) | 0.906 |
| DCO (% predicted) | 82 | -4.949±4.602<br>(-14.114 to 4.217) | 0.285 | 82 | -0.02±0.14<br>(-0.30 to 0.27) | 0.913 |

Footnote: Models were adjusted for age, sex, height, and weight. Abbreviation: FEV<sub>1</sub>, forced expiratory volume in 1 second; FVC, forced vital capacity; FEF, forced expiratory flow; DCO, diffusing capacity; PE, parameter estimate; SEM, standard error of mean; CI, confidence interval.

## DISCUSSION

In this study, we found that never-smoking flight attendants who had a remote history of occupational exposure to SHS had significantly higher systemic (plasma) levels of elastin degradation markers, desmosine and isodesmosine (D/I), compared to nonsmoking individuals with no such history of occupational SHS exposure. Furthermore, we found that among those never-smoking SHS-exposed flight attendants, those with spirometric COPD had higher systemic levels of D/I compared to those without spirometric COPD. While the source of the higher systemic elastin degradation markers in those exposed to cabin SHS (and also those with spirometric COPD) that we observed in this study is unknown, the inverse association of plasma D/I with lung function does suggest that pulmonary elastin degradation at least in part contributes to this process. This is remarkable because it implies that even 25 years after the occupational exposure to cabin SHS (smoking was banned on all domestic and international flights after 1995), there is ongoing differential elastin damage in these never-smokers who were exposed to SHS in the cabin. While there have been previous reports about increased systemic levels of D/I in the setting of acute exposure to SHS,^13^ to our knowledge, this is the first report of describing evidence of lung damage with such remote exposure to SHS.

Many studies have reported D/I levels to be an indicator of lung damage associated with decreased lung function.^14, 38-42^ First, studies that supported the association between destruction of lung elastin with occurrence of airspace enlargement characteristic for emphysema and increased urinary D/I were done in animal models.^39, 40, 42^ Later, human studies confirmed similar findings.^15, 41, 42^ Stone et al extensively explored levels of D/I in relation to presence of COPD, or with smoking status.^43^ They found higher excretion of D/I in urine of COPD patients, and in smokers without presence of COPD compared to healthy lifetime nonsmokers, suggesting smoking and the presence of COPD to be independently associated with elevated urinary D/I levels. Other studies have reported increased D/I levels in smokers and COPD patients, even before clinical symptoms of COPD occurrs.^14^ Slowik et al demonstrated that not only smokers, but also non-smokers exposed to SHS, have higher levels of D/I compared to a control without SHS exposure.^13^ Although, prior studies reported increased plasma D/I due to SHS exposure, our study is the first to document a long-term effect from remote past SHS exposure on systemic levels of D/I, an association between measures of lung function including lung function indices suggestive of small airways and distal lung damage with plasma levels of D/I and past SHS exposure.

Our finding that cabin SHS exposure from many years ago is associated with current elevation of systemic levels of elastin degradation products is quite remarkable but its biological plausibility is supported by the available literature. Excessive and persistent inflammation is a driving force in lung injury and development of COPD, and several studies have shown that airway inflammation persists long after cessation of smoke exposure, including SHS.^44-49^ These findings suggest exposure to smoke could initiate self-perpetuating processes that prevent resolution of the inflammatory response, which in turn cause persistent injury and damage to the lung evident from the elevated levels of elastin degradation products many years after, despite removal of the original insult.

Although no longer being exposed to the high intensity SHS that they experienced in aircraft cabins, flight attendants experience increased rates of respiratory illnesses, compared to the general population.^7-9,3, 7^ McNelly et al^9^ reported roughly three-fold increase in chronic bronchitis prevalence among flight attendants, when compared to age matched general U.S. population in the National Health and Nutrition Examination Survey (NHANES), despite the flight attendants having lower prevalence of smoking. Furthermore, the odds of being diagnosed with chronic bronchitis increased with tenure of flight attendants.^9^ In addition, Beatty et al found increased prevalence of chronic bronchitis, emphysema/COPD, and sinus problems among FAs compared to the U.S. general population.^7^ Arjomandi et al reported never-smoking flight attendants who were exposed to past cabin SHS to have decreased diffusing capacity, with more than half of them having diffusing capacity below the lower limit of the 95% prediction interval for their age, sex and height.^10, 16^ Further, they also had decreased maximal airflow at mid- and low-lung volumes together with pulmonary function evidence of air trapping suggesting airflow obstruction.^3^

## LIMITATIONS

Our study has several limitations. First, exposure to cabin SHS was estimated based on self-reported data regarding airline employment years. Yet, airline employment history could provide a relatively accurate measure of cabin SHS exposure. Moreover, any error in the flight attendants’ recall of employment history is not expected to be related to plasma D/I or lung measurements.

Second, while we observed significant association between history of cabin SHS exposure and higher plasma D/I levels in the univariate linear regression modeling, in the multivariate modeling, years of cabin SHS exposure association with plasma D/I levels did not reach statistical significance after adjusting for age, sex, height, and weight. This could be due to the relationship between D/I levels and years of SHS exposure not being linear; it is possible there is a threshold effect of SHS. Indeed, this is consistent with previous studies reporting the association between diffusing capacity and SHS exposure among those FA who were exposed to SHS for more than 10 years.^10^ Nevertheless, in the current study, in the categorical comparisons of exposed and unexposed participants, plasma D/I levels were significantly higher in those exposed to SHS as a group compared to the unexposed after adjusting for covariates (age, sex, height, and weight). Third, beyond the history of cabin SHS exposure in the past, flight attendants face other environmental exposures, such as low atmospheric pressure and radiation exposure, which could contribute to lung damage measured by D/I levels. However, we observed significant association only between years of cabin SHS exposure and plasma D/I, and did not observe significant association between total years of airline employment and D/I levels.

## INTERPRETATION

Collectively, our study documents the long-term adverse effects of exposure to SHS on pulmonary structure and function. It provides evidence that past exposure to SHS, even when remote, is associated with higher systemic elastin degradation markers, which in turn is indicative of continued lung damage and declining lung function despite the cessation of culprit exposure. Our study furthermore implicates plasma D/I as a sensitive biomarker of lung damage in at risk population regardless of whether their risk of disease is due to current or past exposure to direct or secondhand tobacco smoke.

## TAKE-HOME POINTS

### Study Question

Whether EDM levels are elevated in persons with history of SHS exposure in the past, and whether those levels are associated with reduced lung function.

### Results

Levels of EDM are elevated in persons with SHS exposure in the past; increase in EDM levels is associated with reduced lung function.

### Interpretation

Plasma D/I may prove to be a sensitive biomarker of lung abnormalities in population that was heavily exposed to SHS in the past.

## Data Availability

All data produced in the present study are available upon reasonable request to the authors.

## ABBREVIATIONS

(CF): Cellulose fiber powders
(COPD): Chronic obstructive pulmonary disease
(D/I): Desmosine/isodesmosine
(DCO): Diffusing capacity
(FAs): Flight attendants
(FAMRI): Flight attendant medical research institute
(FEF): Forced expiratory flow
(FEV_1_): Forced expiratory volume in 1 second
(FVC): Forced vital capacity
(SHS): Solid phase extraction
(SPE): Secondhand smoke

## Notes

**Summary conflict of interest:** Authors report no conflict of interest related to this work

**Funding:** This work was supported by: 1. Flight Attendant Medical Research Institute (FAMRI) (012500WG and CIA190001 to MA; CIA150034 to GMT; FAMRI Center of Excellence Award#012500 to RR). 2. California Tobacco-Related Disease Research Program (TRDRP) (T29IR0715 to MA). 3. The Department of Veterans Affairs (CXV-00125 to MA). 4. National Library of Medicine Training Grant (NIH: T15LM007442 to SZ) 5. Postdoctoral Training in Tobacco Control Research (NIH/NCI: T32 CA113710 to JMR) The funders had no role in study design, data collection and analysis, decision to publish, or preparation of the manuscript. The statements and conclusions in this publication are those of the authors and not necessarily those of the funding agency. The mention of commercial products, their source, or their use in connection with the material reported herein is not to be construed as an actual or implied endorsement of such products.

### Competing Interest Statement

The authors have declared no competing interest.

### Funding Statement

This study was supported by:
1.Flight Attendant Medical Research Institute (FAMRI) (012500WG and CIA190001 to MA; CIA150034 to GMT; FAMRI Center of Excellence Award#012500 to RR).
2.California Tobacco-Related Disease Research Program (TRDRP) (T29IR0715 to MA).
3.The Department of Veterans Affairs (CXV-00125 to MA).
4.National Library of Medicine Training Grant (NIH: T15LM007442 to SZ)
5.Postdoctoral Training in Tobacco Control Research (NIH/NCI: T32 CA113710 to JMR)

### Author Declarations

Institutional Review Board (IRB) of the University of Rochester Medical Center (URMC), University of North Carolina (UNC), and University of California, San Francisco (UCSF) gave ethical approval for this work.

