## Supplemental appendix for "Elastin degradation markers are elevated in never-smokers with past history of prolonged exposure to secondhand tobacco smoke and are inversely associated with their lung function": Medrxiv_Suppl_SHS and elastin degradation.docx

**Title:**

**Short Title:** Elastin degradation with secondhand smoke exposure

**Authors:**

Jelena Mustra Rakic, PhD*^,1,2^, Siyang Zeng, MS*^,3,4^, Linnea Rohdin-Bibby^5^, Erin L Van Blarigan, ScD^6^, Xingjian Liu, PhD^7^, Shuren Ma, PhD^7^, John P Kane, MD, PhD^2^, Rita Redberg, MD, MS^5,8^, Gerard M. Turino, MD^7^, Eveline Oestreicher Stock, MD^#,2,5,8^, Mehrdad Arjomandi, MD^#,1,3,9,10^

* These authors contributed equally to this work.

\# Co-senior authors.

^1^ Center for Tobacco Control Research and Education, University of California, San Francisco, California, USA

^2^ Cardiovascular Research Institute, University of California, San Francisco, California, USA

^3^ Medical Service, San Francisco Veterans Affairs Medical Center; San Francisco; California, USA

^4^ Department of Biomedical Informatics and Medical Education, University of Washington, Seattle, Washington, USA

^5^Flight Attendant Medical Research Institute (FAMRI) Bland Lane Center of Excellence on Secondhand Smoke, University of California, San Francisco, California, USA

^6^Department of Epidemiology and Biostatistics, University of California, San Francisco, California, USA

^7^Department of Medicine, Mt Sinai-St Luke's-Roosevelt Hospital, New York, New York, USA

^8^Division of Cardiology, University of California, San Francisco, California, USA

^9^Division of Pulmonary, Critical Care, Allergy and Immunology, and Sleep Medicine, University of California, San Francisco, California, USA

^10^Division of Occupational and Environmental Medicine; University of California, San Francisco, California, USA

^§^**Corresponding author:**

Mehrdad Arjomandi, MD

Department of Medicine

University of California, San Francisco

San Francisco Veterans Affairs Medical Center

Building 203, Room 3A-128, Mailstop 111-D

4150 Clement Street, San Francisco, CA 94121

### **Supplemental Tables**

#### **Table S1**

**Associations of plasma levels of elastin degradation products (desmosine and isodesmosine, D/I) with lung function measures.**

|  | **All particiapnts** | | | **Exposed** | | | **Exposed w/o COPD** | | | **Exposed w COPD** | | |
| --- | --- | --- | --- | --- | --- | --- | --- | --- | --- | --- | --- | --- |
|  | **N** | **PE ± SEM**  **(95% CI)** | **P-value** | **N** | **PE ± SEM**  **(95% CI)** | **P-value** | **N** | **PE ± SEM**  **(95% CI)** | **P value** | **N** | **PE ± SEM**  **(95% CI)** | **P-value** |
| **Airway indices** | | | | | | |  | | |  |  |  |
| FEV_1_ (L) | 288 | -1.76±0.38  (-2.51 to -1.01) | <0.001 | 187 | -1.47±0.46  (-2.36 to -0.57) | 0.001 | 148 | -0.59±0.48  (-1.54 to 0.36) | 0.222 | 39 | -1.88±1.03  (-3.97 to 0.21) | 0.076 |
| FEV_1_ (% predicted) | 288 | -65.84±14.59  (-94.56 to -37.12) | <0.001 | 187 | -57.51±18.35  (-93.71 to -21.3) | 0.002 | 148 | -24.08±20.04  (-63.71 to 15.54) | 0.231 | 39 | -75.89±38.28  (-153.78 to 1.99) | 0.055 |
| FVC (L) | 288 | -1.43±0.46  (-2.34 to -0.52) | 0.002 | 187 | -0.88±0.53  (-1.93 to 0.16) | 0.096 | 148 | -0.62±0.62  (-1.84 to 0.60) | 0.318 | 39 | -1.03±1.24  (-3.56 to 1.49) | 0.412 |
| FVC (% predicted) | 288 | -40.38±13.6  (-67.14 to -13.62) | 0.003 | 187 | -26.79±16.46  (-59.28 to 5.7) | 0.105 | 148 | -20.35±19.80  (-59.48 to 18.79) | 0.305 | 39 | -33.38±35.09  (-104.78 to 38.02) | 0.348 |
| FEV_1_/FVC (%) | 288 | -0.26±0.05  (-0.36 to -0.15) | <0.001 | 187 | -0.31±0.07  (-0.44 to -0.17) | <0.001 | 148 | -0.05±0.05  (-0.14 to 0.05) | 0.341 | 39 | -0.57±0.16  (-0.89 to -0.25) | <0.001 |
| FEV_1_/FVC (% predicted) | 288 | -33.56±6.76  (-46.87 to -20.25) | <0.001 | 187 | -39.92±8.85  (-57.38 to -22.45) | <0.001 | 148 | -5.91±6.05  (-17.87 to 6.06) | 0.330 | 39 | -74.09±20.62  (-116.05 to -32.14) | 0.001 |
| **Small airway indices** | | | | | | |  | | |  |  |  |
| FEF_25-75_ (L/s) | 287 | -2.74±0.61  (-3.94 to -1.54) | <0.001 | 186 | -2.55±0.72  (-3.98 to -1.13) | <0.001 | 147 | -0.98±0.76  (-2.49 to 0.52) | 0.198 | 39 | -1.63±0.82  (-3.3 to 0.03) | 0.054 |
| FEF_25-75_ (% predicted) | 286 | -125.52±27.04  (-178.75 to -72.28) | <0.001 | 185 | -125.05±34.06  (-192.26 to -57.84) | <0.001 | 146 | -51.08±36.28  (-122.80 to 20.65) | 0.161 | 39 | -82.48±39.41  (-162.66 to -2.3) | 0.044 |
| **Distal (parenchymal) lung indices** | | | | | | |  | | |  |  |  |
| DCO (mL/min/mm Hg) | 82 | -9.98±4.71  (-19.37 to -0.59) | 0.037 | 73 | -10.92±4.78  (-20.46 to -1.37) | 0.025 | 62 | -7.48±5.48  (-18.47 to 3.50) | 0.177 | 11 | -12.58±9.72  (-36.36 to 11.2) | 0.243 |
| DCO (% predicted) | 82 | -38.26±18.34  (-74.79 to -1.73) | 0.040 | 73 | -41.81±18.9  (-79.53 to -4.09) | 0.030 | 62 | -28.14±21.68  (-71.57 to 15.30) | 0.199 | 11 | -55.1±38.45  (-149.19 to 38.99) | 0.201 |

Footnote: Regression models were adjusted for age, sex, height, and weight. Abbreviations: FEV_1_, forced expiratory volume in 1 second; FVC, forced vital capacity; FEF, forced expiratory flow; DCO, diffusing capacity; PE, parameter estimate; SEM, standard error of mean; CI, confidence interval. Models were adjusted for age, sex, height, and weight.

#### **Table S2**

**Association of cabin SHS exposure with plasma levels of elastin degradation product (desmosine and isodesmosine, D/I).**

|  | **PE±SEM** | **95% CI** | **P-value** |
| --- | --- | --- | --- |
| **All subjects** | | | |
| **Multivariate (N = 289)** | |  |  |
| Cabin SHS Exposed (Y/N) | 0.0345±0.0092 | 0.0163 to 0.0527 | <0.001 |
| Age (years) | 0.0046±0.0006 | 0.0034 to 0.0057 | <0.001 |
| Sex (F) | 0.0250±0.0106 | 0.0042 to 0.0458 | 0.018 |
| Height (cm) | -0.0006±0.0006 | -0.0018 to 0.0007 | 0.379 |
| Weight (kg) | 0.0007±0.0004 | -0.0001 to 0.0015 | 0.072 |
| **Exposed subgroup** | | | |
| **Univariate (N = 193)** | |  |  |
| Years of SHS exposure (years) | 0.0022±0.0006 | 0.0010 to 0.0034 | <0.001 |
| Years of flight history (years) | -0.0004±0.0005 | -0.0014 to 0.0006 | 0.431 |
| Age (years) | 0.0052±0.0007 | 0.0038 to 0.0066 | <0.001 |
| **Multivariate (N = 187)** | |  |  |
| Years of SHS exposure (years) | 0.0006±0.0009 | -0.0012 to 0.0024 | 0.502 |
| Years of flight history (years) | -0.0008±0.0007 | -0.0021 to 0.0004 | 0.199 |
| Age (years) | 0.0046±0.0009 | 0.0028 to 0.0064 | <0.001 |
| Sex (F) | 0.0225±0.0149 | -0.0069 to 0.0520 | 0.132 |
| Height (cm) | -0.0012±0.0009 | -0.0029 to 0.0005 | 0.160 |
| Weight (kg) | 0.0007±0.0005 | -0.0004 to 0.0018 | 0.215 |

Footnote: Models were adjusted for age, sex, height, and weight. Abbreviation: PE, parameter estimate; SEM, standard error of mean; CI, confidence interval.
